# Impulse dispersion of aerosols during playing the recorder and evaluation of safety measures

**DOI:** 10.1101/2022.04.05.22273294

**Authors:** Marie Köberlein, Laila Hermann, Sophia Gantner, Bogac Tur, Gregor Peters, Caroline Westphalen, Tobias Benthaus, Michael Döllinger, Stefan Kniesburges, Matthias Echternach

## Abstract

**Introduction:** Group musical activities using wind instruments have been restricted during the CoVID19 pandemic due to suspected higher risk of virus transmission. It was presumed that the aerosols exhaled through the tubes while playing would be ejected over larger distances and spread into the room due to jet stream effects. In particular, the soprano recorder is widely used as instrument in school classes and for beginners of all age groups in their musical education, as well as in contexts of leisure activities and in professional concert performances.

Understanding the aerosol impulse dispersion characteristics of playing the soprano recorder could assist with the establishment of concepts for safe music-making.

**Methods:** Five adult professionally trained soprano recorder players (4 female, 1 male) played four bars of the main theme of L. van Beethoven’s “Ode to Joy” in low and in high octaves, as well as with 3 different potential protection devices in the high octave. For comparison they spoke the corresponding text by F. Schiller. Before each task, they inhaled .5 L of vapor from an e-cigarette filled with base liquid. The vapor cloud escaping during speaking or playing was recorded by cameras and its spread was measured as a function of time in the three spatial dimensions. The potential safety devices were rated in practicability with a questionnaire, and their influence on the sound was compared, generating a long-term average spectrum from the audio data.

**Results:** When playing in the high octave, at the end of the task the clouds showed a median distance of 1.06 m to the front and .57 m diameter laterally (maxima: x: 1.35 m and y: .97 m). It was found that the clouds’ expansion values in playing the recorder with and without safety measures are mostly lower when compared to the ordinary, raised speaking voice of the same subjects.

The safety devices which covered the instrument did not show clear advantages and were rated as unpractical by the subjects. The most effective reduction of the cloud was reached when playing into a suction funnel.

**Conclusion:** The aerosol dispersion characteristics of soprano recorders seem comparable to clarinets. The tested safety devices which covered holes of the instrument did not show clear benefits.

## Introduction

During the CoVID19 pandemic, musical activities such as choir singing or wind instrument playing have been suspected of virus transmission via exhaled droplet particles^1,2^, and have therefore been restricted by laws in many countries around the world. The combination of jet stream effects, accumulation of exhaled particles and group gatherings led to the closing of concert halls, as well as the prohibition of musical events, rehearsals, and music lessons in schools^3^. Therefore, institutions in which playing a wind instrument plays an important role, such as theatres, concert halls, schools, music schools and music universities had to invent safety concepts to be able to maintain musical activities. These concepts often included frequent disinfection of the room, frequent aeration and distancing or even teaching completely online, as well as covering the instruments with masks or fabrics^4–10^. In order to provide helpful information for the development of such concepts, several studies tried to figure out if wind instruments were increasing the infection risks due to both aerosol generation and dispersion, or if the instruments were decreasing the risks by capturing the condensed aerosols inside the instruments. While a study on vuvuzelas, which are straight narrow tubes comparable to recorders, showed the ejection of much higher amounts of aerosols than in shouting^11^, a more recent study found smaller aerosol expulsion from classical wind instruments compared to vocalization^12^. Another study divided various orchestral wind instruments into different risk categories by their aerosol emissions^13^. They found flutes and clarinets, which resemble the recorder in mechanism and shape, respectively, to be only at intermediate transmission risk, while trumpet and oboe belonged to the high-risk category. In contrast, a study analyzing the impulse dispersion of aerosols ranked the flute to be more risky than clarinet and trumpet, due to the larger distances reached by the aerosols^14^. A study by Stockman et al.^15^ found comparable amounts of airborne particles in clarinets and singing, as well as effective reduction of aerosol concentration and dispersion by surgical masks. Nevertheless, their results showed high unsteadiness and situation-dependent variations in the concentrations and dispersions of the emitted vapor clouds.

All these studies concentrated on a variety of orchestral instruments, while the recorder, which is a common instrument for musical education and leisure activity in children and adult amateurs as well as professional musicians, has, to the best of the authors’ knowledge, not been addressed, and it could be speculated that the results of other studies cannot be transferred simply. Referring to a representative study on amateur music in Germany, the recorder is played by 22% of female and 3% of male amateur musicians aged 16+. In children aged 6-15, 96% play an instrument, with the recorder found in the second place by popularity, being played by 24% of these children^16^. The soprano recorder is, furthermore, used in music classes of public schools all over the world for its low purchase costs and its ease of use and, consequently, the actual number of children using recorders, even if not considered as a musical hobby, can be assumed to be much higher.

In order to help facilitate the risk management and find suitable measures for recorder classes, the presented study investigates the properties of the impulse dispersion of aerosol clouds emitted during playing soprano recorders. It also evaluates several protective devices that had been invented with the expectation of decreasing the radius of aerosol dispersion.

## Material and Methods

After the approval of the local ethical committee (20-1065), five full-time student recorder players from the local University of Music (4 female, 1 male) were recruited. None of the subjects showed respiratory problems in their respective medical histories and spirometry (ZAN, Inspire Healthcare, Oberthulba, Germany). There were no signs of acute infection based on their answers to an acute infection questionnaire.

### Tasks and setup

In the experiment, the subjects performed six tasks (for access to Figure 1 please contact the corresponding author):

1. Speaking the sentence “Freude schöner Götterfunken, Tochter aus Elisium” (“Ode to joy” by Friedrich Schiller (used also by Beethoven in his ninth symphony, 4^th^ movement)) with raised voice.
2. Loudly playing four bars of the melody of the “Ode to joy” by Ludwig van Beethoven in their lower octave, starting from F#5 (fundamental frequency (*f*_o_) approx. 740 Hz). The speed was set as approx. 120 bpm.
3. Loudly playing the same melody in their higher octave, starting from F#6 (*f*_o_ approx. 1475 Hz).
4. Playing the same melody in their higher octave with a specially designed extended cloth-mask that covered the player’s mouth and nose and the entire instrument (material: 200g/m2 cotton molton), as well as a cover tissue for the bell hole called “Plopp-Schutz” (©Bonner Textilmanufaktur^17^).
5. Playing the melody in the higher octave, the bell hole covered with household paper towel, as suggested by Bauhaus-Universität Weimar^5^.
6. Playing the above-mentioned melody in the higher octave into a suction funnel (diameter at the inlet: .6 m, diameter at the outlet: .1 m, length .45 m, extended by a pipe of 1 m; operated by a usual axial fan obtained from IT hardware).

**Fig. 1.**
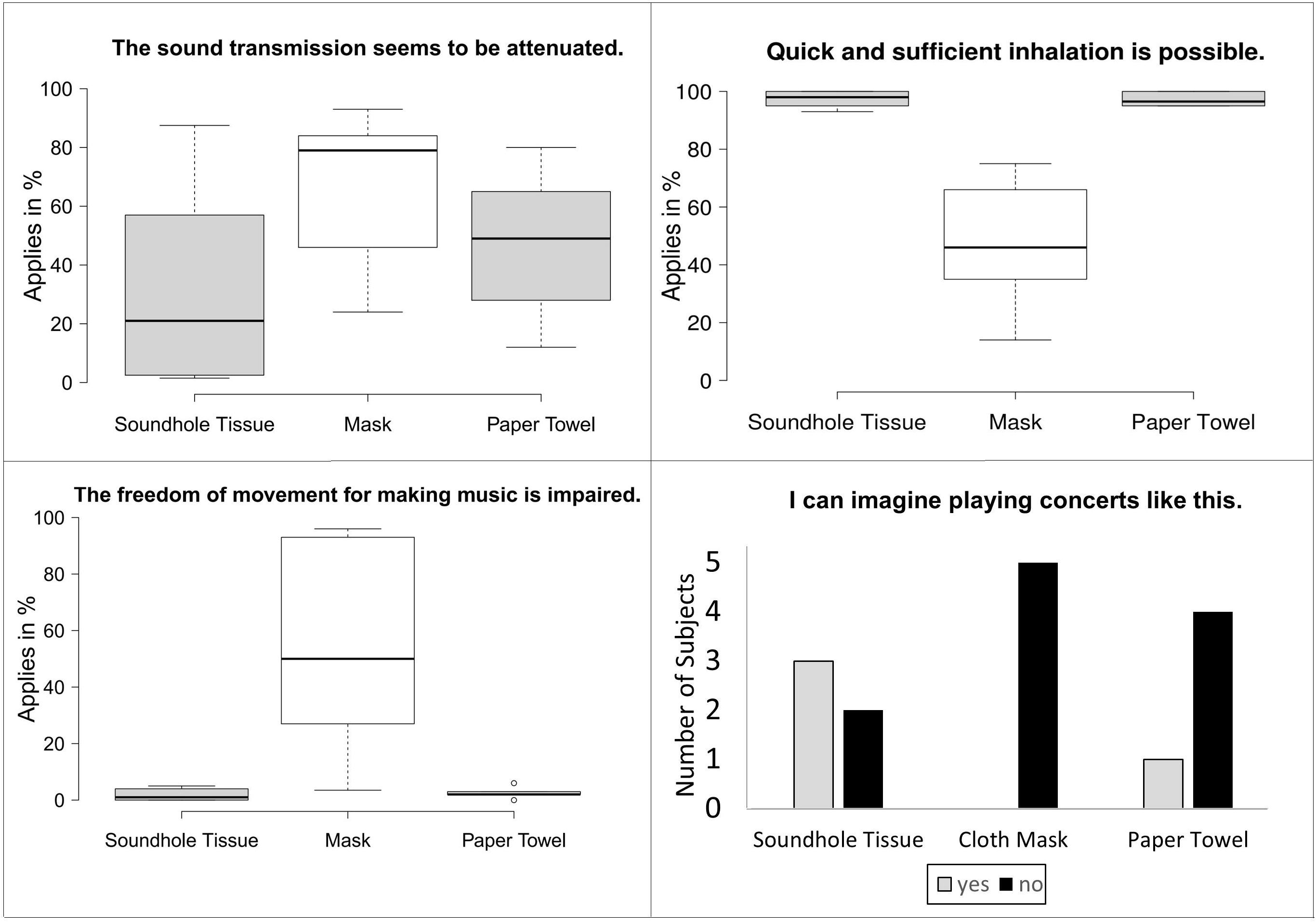
All performed tasks: (1) Speaking the quote from Friedrich Schiller’s “Ode to Joy”^27^. (2) and (3) playing the melody. The potential protection devices: (4) cloth mask and bell hole cover, (5) paper towel, (6) suction funnel.

Each task lasted approximately 8 seconds. Before the subjects started the tasks, they inhaled .5 l vapor from the basic liquid through a Lynden Vox e-cigarette (Lynden GmbH). The basic liquid consists of 50% glycerin and 50% polypropylene glycol. The aerosols in the e-cigarette vapor have the size in the range of 250-450 nm, which approximately matches the size of aerosols generated by human expiration^18^. A spirometer (ZAN, Inspire Healthcare, Oberthulba, Germany) was mounted on top of the e-cigarette to measurethe volume of the vapor inhaled by the subject.

To avoid convectional flows, the subjects were standing on a marked point on a stage while inhaling and performing. They were asked to avoid any motion after finishing the tasks. As in previous experiments^14,19–23^, the measurements were conducted in a Bavarian Television Broadcasting Studio with the dimensions of 27 m x 22 m x 9 m. Three synchronized high-definition television cameras (Sony, Tokio, Japan, resolution 1920 × 1080 pixels, 25 fps) recorded the tasks from a side view (C1), a front view (C2) and a top view (C3). The studio was aerated before and after each task with opposed doors open for a minimum of 2 minutes. The room had a constant humidity of 29.8% and a constant temperature of 21.0°C. Three spotlights illuminated the white aerosol clouds against the black studio walls. For the conversion of pixels into metric values in the data processing, 3 metric scaling bars were installed at the performance stage to cover all 3 spatial directions.

### Data analysis

Similar to previous experiments^14,19–23^, the video footage was converted into negative black and white. The subjects were asked to wear black clothes. However, some uncovered parts of the human body like hands or faces were masked to avoid segmentation errors. The expansions of the expelled aerosol clouds were segmented in each video frame using the software Glottis Analysis Tools (University Erlangen-Nürnberg, Germany)^24^. The footage of C2 had to be excluded from further analysis due to light reflections which led to artifacts in the segmentation process. The footage of C1 was used to measure the maximum expansion to the subjects’ front (x-direction) and in maximum positive and negative vertical direction (z-direction). The footage of C3 provided the side expansion of the aerosol clouds (for access to Figure 2 please contact the corresponding author). The position of the mouth was set as starting point of the aerosol cloud dispersion.

**Fig. 2:**
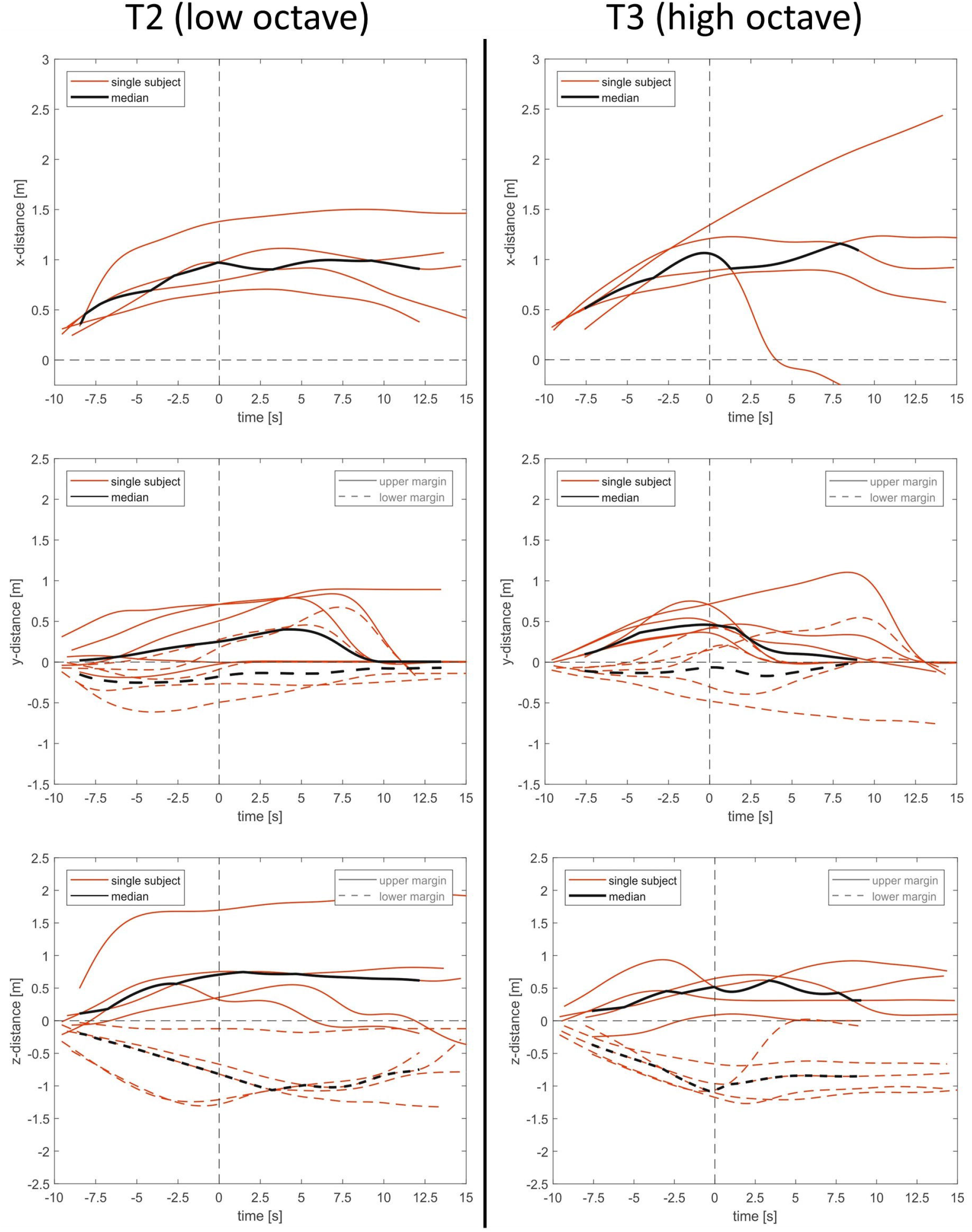
Pictures of camera C1 (left) recording the x-dimension to the front and the vertical z-dimension from a side view, and camera C3 (right) recording the y-dimension to the sides from a top view.

For the temporal representation of the clouds’ dispersion, the time point zero was defined as the end of a task (0 point in the time domain), i.e., where phonation or playing stopped. Consecutively, the start of the task had negative values in the time domain.

The temporal devolution of the maximum dispersion in each direction was filtered by a moving median filter (window length of 30 time points) for each player and subsequently smoothed by a cubic spline interpolation in order to remove segmentation outliers, both steps carried out in Matlab (The Mathworks Inc., Natick, MA).

In task 6 (suction funnel), the pipe’s end was not connected to a longer tube which would guide the vapor outside of the room so that the vapor was released into the studio. However, this released cloud was not considered in the segmentation as it was clearly distinguishable from the clouds directly originating from the player and/or the recorder.

### Acoustical evaluation of the potential safety device

Additionally, the subjects were asked to perform the melody of task 3 without e-cigarette vapor but in 30 cm distance to a D:SCREET 4061 microphone (DPA Microphones A/S, Alleroed, Denmark). The microphone was connected to the computer via a Scarlett 2i2 audio interface (Focusrite Plc, High Wycombe, England). The audio signal was then sampled with the software Audacity26 (MuseCY SM Ltd, Limassol, Cyprus) at a sampling rate of 44.1 kHz. The melody was performed (1) with each of the potential safety devices and (2) without any. From the audio recordings, Long Term Average Spectra (LTAS) were obtained using the software Wavesurfer (version 1.8.8, Centre for Speech Technology, KTH Stockholm; settings: FFT: Hanning, 512 points, range: -110 dB), in order to detect changes in the spectral characteristic of the instrument’s sound caused by the safety measures.

### Subjective evaluation of the potential safety device

The subjects also answered a questionnaire consisting of visual analogue scales (0-100 mm) with statements on the freedom of movement, sound attenuation, inhalation sufficiency, and concert practicability to subjectively evaluate the application of the analyzed safety devices, see supplement.

Due to the small number of subjects, statistical calculations were considered not meaningful.

## Results

All subjects were able to perform all tasks without any dropout. The vapor escaped the instrument mainly through the labium and the bell hole and merged into one cloud.

At the end of the speaking task, the maximum expansion of the aerosol clouds reached 1.32 m to the front, with a median of .83 m. The dispersion at the end of the task for playing the recorder in the low and the high octave exhibited maximum values comparable to speaking, i. e., in a range of ± .1 m, with slightly higher median values to the front (T2: 0.97 m; T3: 1.06 m), see table 1.

**Table 1.** The maximum and median values for all performed tasks at the time point 0 s (end of the task), as well as 3 s and 10 s after the end of the task. *Italic* values in T2-3 show an at least .1 m higher value in comparison to T1. **Bold values** are more than .1 m below the compared value from T1. T4-6 are compared to T3: Bold values are at least .1 m below the compared value of T3, narrow markings show an at least .1 m higher value than in T3. Unmarked values are within a ± .1 m range of the compared value.

| Speaking T1 |  |  |  |  |  |  |
| --- | --- | --- | --- | --- | --- | --- |
|  | x-Dimension |  |  | Diameter y-Dimension |  |  |
|  | 0[s] | 3[s] | 10[s] | 0[s] | 3[s] | 10[s] |
| Median | 0.83 m | 1.12 m | 1.41 m | 0.84 m | 0.97 m | 0.58 m |
| Maximum | 1.32 m | 1.89 m | 2.58 m | 1.16 m | 1.43 m | 1.62 m |
| Playing Low Octave T2 |  |  |  |  |  |  |
|  | x-Dimension |  |  | Diameter y-Dimension |  |  |
|  | 0[s] | 3[s] | 10[s] | 0[s] | 3[s] | 10[s] |
| Median | *0.97 m* | **0.90 m** | **0.97 m** | **0.47 m** | **0.42 m** | **0.04 m** |
| Maximum | 1.38 m | **1.44 m** | **1.50 m** | **0.77 m** | **1.00 m** | **1.14 m** |
| Playing High Octave T3 |  |  |  |  |  |  |
|  | x-Dimension |  |  | Diameter y-Dimension |  |  |
|  | 0[s] | 3[s] | 10[s] | 0[s] | 3[s] | 10[s] |
| Median | *1.06 m* | **0.93 m** | **1.01 m** | **0.57 m** | **0.53 m** | **0.23 m** |
| Maximum | 1.35 m | **1.62 m** | **2.17 m** | **0.97 m** | **0.83 m** | **0.72 m** |
| Mask and Bell Hole Tissue (T4) |  |  |  |  |  |  |
|  | x-Dimension |  |  | Diameter y-Dimension |  |  |
|  | 0[s] | 3[s] | 10[s] | 0[s] | 3[s] | 10[s] |
| Median | **0.65 m** | **0.81 m** | **0.70 m** | 0.56 m | *0.67 m* | *0.65 m* |
| Maximum | 1.40 m | *1.74 m* | *2.28 m* | **0.78 m** | 0.77 m | 0.81 m |
| Paper Towel (T5) |  |  |  |  |  |  |
|  | x-Dimension |  |  | Diameter y-Dimension |  |  |
|  | 0[s] | 3[s] | 10[s] | 0[s] | 3[s] | 10[s] |
| Median | *1.17 m* | *1.28 m* | *1.36 m* | 0.59 m | *0.70 m* | *0.70 m* |
| Maximum | 1.31 m | **1.51 m** | **1.93 m** | **0.74 m** | *0.96 m* | *1.29 m* |
| Suction Funnel (T6) |  |  |  |  |  |  |
|  | x-Dimension |  |  | Diameter y-Dimension |  |  |
|  | 0[s] | 3[s] | 10[s] | 0[s] | 3[s] | 10[s] |
| Median | **0.65 m** | **0.42 m** | **0.39 m** | **0.19 m** | **0.08 m** | **0.08 m** |
| Maximum | **1.16 m** | **1.14 m** | **1.46 m** | **0.65 m** | **0.55 m** | **0.62 m** |

The side expansion of the clouds from speaking at the end of the task showed a maximum diameter of 1.16 m with a median of .84 m. All values of the aerosol clouds’ lateral expansions during playing the recorder in high and low octave were lower than in speaking (table 1).

At the time points 3 and 10 seconds after the end of the task, the clouds’ expansions from playing the recorder were found below all compared values of the speaking task. A maximum expansion to the front of 2.17 m was reached by one subject 10 seconds after playing in the high octave, which is still .41 m below the maximum value 10 seconds after speaking (2.58m).

A maximum side expansion diameter of 1.14 m was reached 10 seconds after playing in the low octave, which is .48 m below the maximum value 10 seconds after speaking (1.62 m).

Figure 3 shows the dispersion for all subjects’ tasks 2 and 3 and the corresponding medians.

**Fig. 3.**
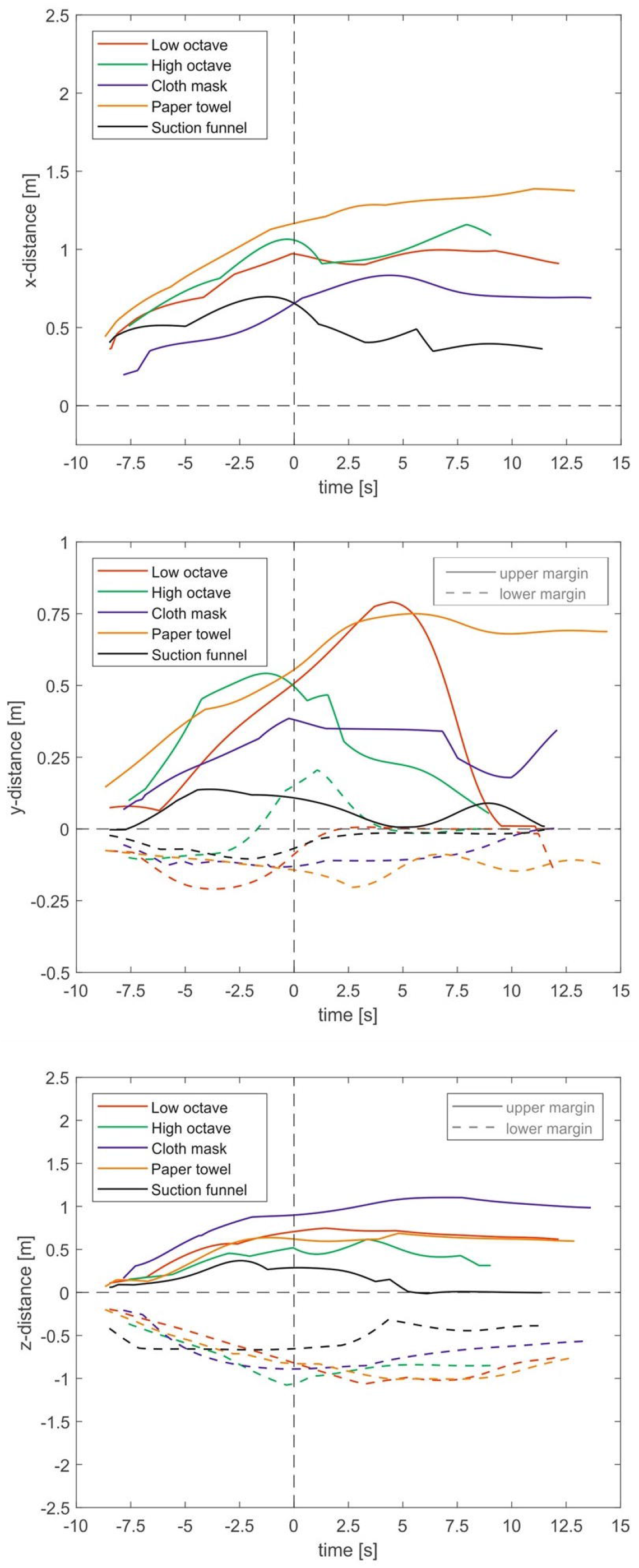
Aerosol dispersion distances of all subjects (red) and medians (black) over all subjects in task 2 (low octave) and task 3 (high octave) for x- (front), y- (side), and z- (vertical) direction. The distances are measured from the subjects’ mouth. The zero in the time domain is set as the end of the task.

All safety devices influenced the dispersion patterns during playing the recorder when compared to task 3, i. e., playing in the high octave without safety device.

When playing with the mask and bell hole cover (T4), the median in x-direction to the front at the end of the task was .41 m below the compared value for task 3 and stayed below the median values for the later time points.

The maximum value to the front at the end of the task was comparable to T3 and 10 seconds later the maximum exceeded the compared value of T3 by .11 m.

With regard to the lateral expansion at the end of the task, the median diameters of T3 and T4 were comparable. The maximum value for T4 at the end of the task was .19 m lower than in T3 (table I).

For task 5, i. e., applying the paper towel during playing, the median values to the front were higher than in T3, while the maxima were below the compared values, but only .04 m lower at the end of the task.

In the y-dimension at the end of the task, the median value was comparable to the value of T3, with a maximum of .23 m below the value of T3. All values to the sides at later time points were higher than in the compared task T3, reaching a largest diameter of 1.29 m after 10 seconds.

All listed values in task 6 with the suction funnel were below the values of T3 (table I).

Figure 4 provides the medians for the safety devices in comparison to normal playing in low and high octave.

**Fig. 4.**
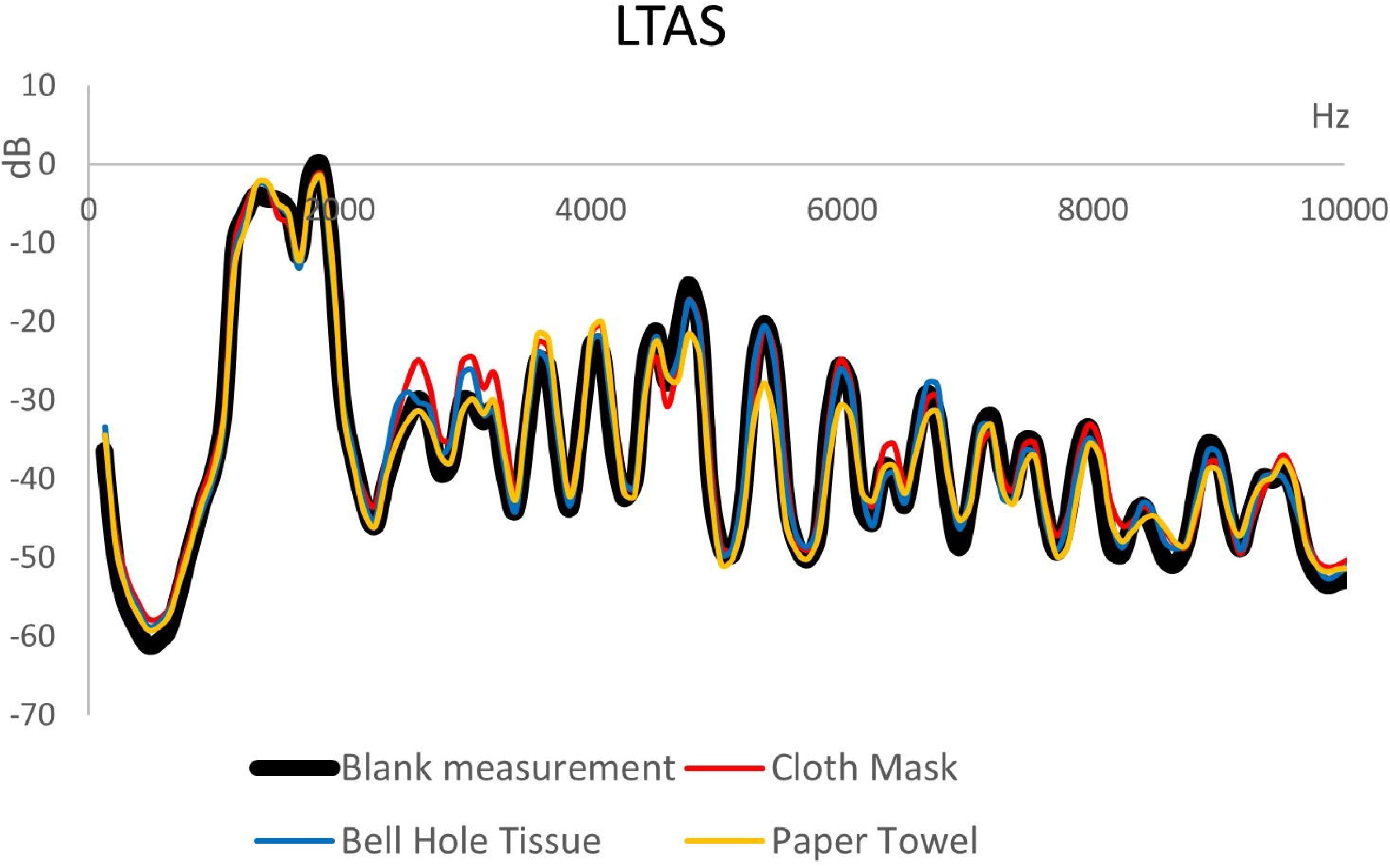
Development of the medians of all three dimensions over time. The zero-point in the time domain marks the end of the phonation task. The distances are measured from the subjects’ mouths.

Regarding the vertical direction, playing in the low octave reached the highest maxima upwards from the subject’s mouth with 1.7 m at the end of the task and 1.91 m at time point 10 s.

The highest median values of all tasks in positive z-direction (upwards) were reached when playing with mask and bell hole tissue, showing .9 m at the end of the task and 1.04 m at time point 10 s.

### Evaluation of safety devices

The long-term average spectrum of playing in the high octave without device (pure) and with the mask or tissue or paper towel is depicted in Figure 5. No significant attenuations or deviations were observed. The mean squared errors are 4.7 for the extended mask, 2.77 for the bell hole tissue and 5 for the paper towel.

**Fig. 5.** Long-term average spectrum of the recorder when playing the same melody with and without potential safety devices.

Figure 6 shows the results of the questionnaires concerning mask, tissue, and paper towel. The perception of sound attenuation was reported for all of the safety devices, but least for the bell hole tissue. For the paper towel, it was shown that the covering of the bell hole prohibited the production of low pitches where all fingers close their assigned hole. Especially for the mask, the musicians reported problems in sufficient inhalation and restriction of movement. All the subjects rejected the application of masks for use in concerts. All except one musician rejected the paper towel. The tissue was rated as practicable in concerts by three of the five subjects.

**Fig. 6.** The subjects’ evaluation of the practicability of the potential safety procedures.

## Discussion

This study investigates the impulse dispersion of aerosols during playing the soprano recorder and three potential safety devices. In general, it was found that the impulse dispersion characteristics were comparable to those of the clarinets^14^ but not flutes, and that a suction funnel can reduce the aerosol dispersion significantly, while other devices covering the instrument were problematic concerning the aerosol dispersion and/or practical implementation.

Group musical activities were restricted in many countries during the Covid19 pandemic, due to the estimated higher risk of virus transmission in playing wind instruments^1,2^. The risk is assumed to be higher due to jet streams through narrow instruments, as well as accumulation and suspension of aerosols in rooms with group gathering.

Aerosol emission rates and impulse dispersion of several orchestra instruments have already been investigated in previous studies^11-15^; however, none of these studies included the recorder, which is a very common instrument, not only in professional settings but also in music education and amateur music-making. Understanding the dispersion characteristics could help in risk management and adjustment of safety concepts.

When playing in the high octave, at the end of the task, the clouds showed medians of 1.06 m to the front (x-direction) and .57 m diameter laterally (y-direction), with maxima of x: 1.35 m and y: .97 m). Compared to the dispersion values of other instruments in a previous study with the same experimental setup^14^, without safety device at the end of the task, the straight-built recorder produced larger distances than the winding-built trumpet (the trumpet values were medians: x: .86 m and y: .3 m; maxima: x: 1.20 m and y: .52 m). Even though the mechanism of the recorder is somewhat similar to the flute, dividing the airflow from the windway above the block at the labium, the generated jet stream of aerosols did not go as far as for the flutes (flutes’ medians: x: 1.51 m, y: .64 m; maxima: x: 1.88 m, y: 1.23 m). The values of the recorder were rather comparable to the shape-resembling clarinet (medians x: 1 m, y: .55 m; maxima x: 1.51 m, y: .95 m).

Similar to the clarinets in the previous study^14^, the clouds’ frontal expansions of the recorders generally stagnated or decreased after the end of the task at a distance of around 1 m. For a single subject the cloud continued to expand and reached a maximum distance of 2.17 m to the front, 10 seconds after playing in the high octave, which exceeds former recommendations of a safety distance of 2 m to the front for wind instruments in general. Accordingly, the lateral expansions decreased after the end of the task and increased for one subject after playing in the low octave, reaching a maximum diameter of 1.14 m 10 seconds after the end of the task. Considering the individually different expansion dimensions of each cloud after the end of the task (see for example fig. 3: T3 in x-direction), it seems likely that, after the primary impulse, multifactorial influences such as convectional flows or temperature differences make the diffusion of the clouds hardly calculable and consequently questionable.

It must be taken into account that this study measured only trained, adult recorder players playing loudly, and that children and amateurs might have less exhalation power and playing at moderate volume was not examined. This might influence the distance of the impulse dispersion.

It should also be noted here that the aerosol dispersion reflects only a part of the potential virus transmission pathway which includes (1) the aerosol generation, (2) the aerosol dispersion, (3) the accumulation in closed rooms, and (4) the inhalation of aerosols by the recipient^25^. No aerosol concentrations were measured in the presented study. However, the mechanism and construction of the recorder could be assumed comparable to flutes and clarinets. Consequently, in application of the data by He et al.^26^ also the recorder could be classified into an intermediate risk category. However, this assumption should be verified in future investigations.

The installation of the extended mask and bell hole tissue, covering all holes of the instrument, did not show a clear pattern in comparison to the task without mask (T3), with values evenly distributed to higher, lower, and comparable categories. Although the median values to the front were lower with the safety procedure, the maxima were higher, reaching 2.28 m after ten seconds demonstrating a greater variance among all measurements.

Playing with a paper towel closing the bell hole tightly showed many values that were even higher than without it, i. e., 9 out of 12 values in x- and y direction (see table I). This might be due to pressure-supercompensation to the perceived resistance at the end of the tube, augmenting the jet stream at the narrow labium. The maximum side diameter of the cloud reached 1.29 m 10 seconds after the end of the task. Consequently, a covering of the bell hole without covering the labium is considered worse than playing without any device.

The suction funnel was the only safety installation that showed convincing effects in all directions at all time points. However, the device needs significant technical effort to be installed correctly while producing as little noise as possible so as not to disturb the music, which could make teachers and institutions hesitant.

Apart from the aerosol dispersion, the subjects did not approve most of the safety devices. The single exception was for the bell hole tissue, with which three of the five subjects stated they could imagine playing a concert. The mask was estimated to impair the free movement and sufficient inhalation for playing. Perceived sound attenuation, including difficulties in intonation, was present more or less for all devices that covered the bell hole. However, attenuation was not visible in the LTAS of any of the installations. Additionally, a tight covering of the bell hole prohibited the playing of low pitches where all finger holes are closed.

Thus, none of the evaluated potential safety measures fulfilled all the needs of the recorder players and the function as protection mechanism.

## Limitations

The presented study concentrated on the impulse dispersion of a single musical phrase. However, in real life situations, the rehearsal of a whole piece takes much longer and, consequently, aerosol clouds could accumulate in the room. Additionally, the study measured motionless, single players. It can be expected that children playing in larger groups will produce flows in the room which would be difficult to predict, which underlines the importance to aerate the room frequently or with opposed windows open.

Furthermore, the measured values represent the distance reached by the aerosol cloud but not the number of aerosols or their concentration in the cloud. Additionally, the potential virus dose in such a cloud differs by subject and in the course of disease.

Another limitation of this study is the rather small sample size of 5 subjects.

## Conclusion

This study investigated the impulse dispersion of aerosols in soprano recorders and evaluated three different potential safety devices concerning the spread of aerosols, i. e., different types of covering of the holes and a suction funnel. The pure aerosol cloud dispersion were found to be comparable to those of the clarinet measured in a previous study. Concerning the evaluated potential safety installations, the different types of covering the instrument did not show clear benefits in prohibiting the expansion of the aerosol clouds and were furthermore rated as impracticable by the subjects. The suction funnel showed almost total reduction of the aerosol clouds and, thus, seems to be a effective method to remove the aerosol particles immediately after the emission from the instrument.

## Data Availability

The datasets generated during and/or analyzed during the current study are available from the repository https://doi.org/10.5281/zenodo.6395624 or from the corresponding author on request.

## Acknowledgements

This study was funded by the Ministry of Science and Art of the State of Bavaria (Germany). The authors thank all members of the Bavarian Broadcast for their help in realizing this study.

## Notes

### Competing Interest Statement

The authors have declared no competing interest.

### Author Declarations

Ethikkommission bei der Medizinischen Fakultaet der LMU Muenchen (Ethics committee of the medical faculty of the university hospital of Munich) gave ethical approval for this work (Votum 20-1065)

